# Trajectories of hospitalisation for patients infected with SARS-CoV-2 variant B.1.1.7 in Norway, December 2020 – April 2021

**DOI:** 10.1101/2021.06.28.21259380

**Authors:** Robert Whittaker, Anja Bråthen Kristofferson, Elina Seppälä, Beatriz Valcarcel Salamanca, Lamprini Veneti, Margrethe Larsdatter Storm, Håkon Bøås, Nina Aasand, Umaer Naseer, Karoline Bragstad, Olav Hungnes, Reidar Kvåle, Karan Golestani, Siri Feruglio, Line Vold, Karin Nygård, Eirik Alnes Buanes

**Affiliations:** Department of Infection Control and Vaccines, Norwegian Institute of Public Health, Lovisenberggata 8, 0456, Oslo, Norway; Department of Method Development and Analytics, Norwegian Institute of Public Health, Lovisenberggata 8, 0456, Oslo, Norway; European Programme for Intervention Epidemiology Training (EPIET), European Centre for Disease Prevention and Control (ECDC), Gustav III:s Boulevard 40 169 73 Solna, Sweden; Department of Infection Control and Preparedness, Norwegian Institute of Public Health, Lovisenberggata 8, 0456, Oslo, Norway; Department of Infectious Disease Registries, Norwegian Institute of Public Health, Lovisenberggata 8, 0456, Oslo, Norway; Department of Virology, Norwegian Institute of Public Health, Lovisenberggata 8, 0456, Oslo, Norway; Department of Anaesthesia and Intensive Care, Haukeland University Hospital, Bergen, Norway; Department of Clinical Medicine, University of Bergen, Bergen, Norway; Norwegian Intensive Care and Pandemic Registry, Haukeland University Hospital, Bergen, Norway

**Keywords:** Norway, SARS-CoV-2, hospitalisation, intensive care, variants of concern

## Abstract

**Background:** The SARS-CoV-2 variant of concern (VOC) B.1.1.7 has spread worldwide and has been associated with increased risk of severe disease. Studies on patient trajectories and outcomes among hospitalised patients infected with B.1.1.7 are essential for hospital capacity planning.

**Methods:** Using linked individual-level data from national registries, we conducted a cohort study on cases of SARS-CoV-2 in Norway hospitalised between 21 December 2020 and 25 April 2021. We calculated adjusted hazard ratios using survival analysis to examine the association between B.1.1.7 and time from symptom onset to hospitalisation, and length of stay (LoS) in hospital and an intensive care unit compared to non-VOC. We calculated adjusted odds ratios using logistic regression to examine the association between B.1.1.7 and mortality (up to 30 days post discharge) compared to non-VOC.

**Results:** We included 946 B.1.1.7 patients and 157 non-VOC. The crude median time from symptom onset to hospitalisation was 8 days (IQR: 5–10) for B.1.1.7 and 8 days (IQR: 4–11) for non-VOC. The crude median LoS in hospital was 5.0 days (IQR: 2.6–10.0) for B.1.1.7 patients and 5.1 days (IQR: 2.5–9.9) for non-VOC. Fifty-four (6%) B.1.1.7 patients died, compared to 14 (9%) non-VOC. There was no difference in the unadjusted or adjusted estimates of our outcome measures for B.1.1.7 and non-VOC patients.

**Conclusions:** B.1.1.7 does not appear to influence hospitalised patient trajectories, compared to non-VOC. These findings, along with the success of ongoing vaccination programmes, are encouraging for ongoing capacity planning in the hospital sector.

## BACKGROUND

The emergence and spread of variants of concern (VOC) of the SARS-CoV-2 virus have been reported worldwide. This includes lineage B.1.1.7 (alpha variant), first detected in south-east England in September 2020 (1). In Norway (population 5.4 million), testing activity for COVID-19 is high, with consistently over 100,000 persons tested weekly (defined as one or more tests per person within a seven-day period) since week 44, 2020. Mathematical modelling has estimated that consistently over 50% of all cases weekly have been diagnosed since late 2020 (2). Sequencing capacity in Norwegian laboratories was rapidly scaled up from early December 2020 and the capacity to screen for variants or perform whole genome sequencing (WGS) was further increased following reports of widespread transmission of B.1.1.7 in the United Kingdom (UK). The proportion of confirmed COVID-19 cases nationally with known variant increased from 6% in week 52 of 2020 to 80% in week 9 of 2021. The first infection with B.1.1.7 was sampled in week 48, 2020, and it has been the dominating variant since week 7, 2021 (2).

In addition to increased transmissibility (1), a few studies have indicated that B.1.1.7 infection is associated with increased risk of hospitalisation, admission to an intensive care unit (ICU) and/or death (3-10). In Norway, increasing detection of B.1.1.7 in early 2021 coincided with a rapid increase in the number of new admissions to hospital and ICU (2). Infection with B.1.1.7 was subsequently associated with a 1.9-fold increased risk of hospitalisation and a 1.8-fold increased risk of ICU admission, compared to non-VOC. However, when the analysis was restricted to hospitalised cases, there was no difference in the risk of ICU admission between B.1.1.7 and non-VOC (11).

To further explore this finding, and support capacity planning in the health system, evidence on differences in patient trajectories and outcomes among hospitalised patients infected with B.1.1.7 compared to other lineages is essential. We used linked individual-level data to estimate the time from symptom onset to hospitalisation, length of stay (LoS) in hospital and ICU, and odds of mortality (in-hospital and post discharge) for infections with B.1.1.7 in Norway, compared to non-VOC.

## METHODS

### The national emergency preparedness register

The emergency preparedness register, Beredt C19, (12) contains individual-level data from central health registries, national registries and national clinical registries. We included data on notified cases of laboratory-confirmed SARS-CoV-2 infection from the Norwegian Surveillance System for Communicable Diseases (MSIS). We obtained data on hospitalisation and intensive care admission following a positive SARS-CoV-2 test from the Norwegian Intensive Care and Pandemic Registry (NIPaR). All Norwegian hospitals report to NIPaR, and reporting is mandatory. Data on virus variants came from the MSIS laboratory database, which receives SARS-CoV-2 test results from all Norwegian microbiology laboratories. Data on COVID-19 vaccinations came from the Norwegian Immunisation Registry, SYSVAK. More detailed information on each data source can be found in supplementary materials A, part 1 and at (12).

### Study design and setting

We conducted a cohort study, including notified cases of COVID-19 who were hospitalised not more than two days before and less than 28 days after a positive SARS-CoV-2 test in Norway between 21 December 2020 and 25 April 2021, who had available variant data after WGS or PCR screening, and who had not been vaccinated with a COVID-19 vaccine before sampling or hospitalisation. We extracted data from Beredt C19 on 2 June 2021, ensuring a minimum of 36 days follow-up since last date of hospital admission.

### Laboratory investigations and classification of variants

Variants were identified based on WGS using Illumina or Nanopore technology, partial sequencing by Sanger sequencing or PCR screening for selected targets. PCR screening methods include real-time RT-PCR, sometimes in combination with melting curve analysis, of one or several mutation targets specific to the different variants. Other VOC (B.1.351, P.1 and B.1.617.2) as defined by the European Centre for Disease Prevention and Control on 24 May 2021 (13), and cases for whom VOC and non-VOC could not clearly be distinguished were excluded. All other variants were classified as non-VOC. Cases of B.1.1.7 and non-VOC are henceforth referred to as our study cohort.

### Outcome measures

We calculated the number of days between symptom onset and hospitalisation using the reported date of symptom onset in MSIS, and time of first admission in NIPaR. We assembled patient trajectories based on data from NIPaR. We calculated the LoS in hospital and ICU as the time between first admission and last discharge. For patients with more than one registered stay in hospital, we included time between stays as part of the patient’s LoS, if the time between two consecutive stays was less than 24 hours, to allow for transfers between hospitals. A similar 12-hour definition was applied to time in ICU. The time in hospital for patients needing ICU treatment was divided in three; time before admission to ICU, time in ICU and time after discharge from ICU. Patients with unknown date of discharge from their last stay were considered to still be admitted to hospital, and patients who additionally had an unknown date of discharge from ICU to still be admitted to ICU. Death was defined as those who died in-hospital and up to 30 days post discharge (see supplementary materials A, part 1).

### Data analysis

We described cases in terms of demographic characteristics, underlying risk factors, microbiological characteristics, hospital and ICU admission, and mortality. We assessed the representativeness of our study population by comparing the characteristics of our study cohort and notified patients using chi-square tests, Wilcoxon rank-sum tests and hazard ratios (HR) (supplementary materials A, part 2.1).

We conducted a similar analysis to assess the representativeness of patients with known date of symptom onset, compared to our study cohort (supplementary materials A, part 2.2). We used Wilcoxon rank-sum tests to test differences in the distribution of time between sampling date and date of first hospitalisation between B.1.1.7 and non-VOC.

Explanatory variables used to analyse differences in our outcomes were virus variant (B.1.1.7 or non-VOC), age (continuous variable either linearly or with a spline), sex, county of residence, regional health authority, week of admission, country of birth (Norway, overseas and unknown), main cause of hospitalisation (COVID-19, other, unknown) and underlying risk factors.

To analyse differences in the time between symptom onset and hospitalisation, and LoS in hospital and ICU, we used survival analysis, with right censoring of those still admitted to hospital and/or ICU at the end of the study period, or that died while admitted. Kaplan Meier curves were computed for each explanatory variable univariably, using survfit from the R-package survival. One minus the empirical cumulative negative binomial distribution function was fitted to each Kaplan Meier curve by minimising the sum of squared error, using the function optim in R. The function coxph from the R-package survival was used to compute the HR for each explanatory variable.

We used logistic regression to estimate the differences in the proportion of patients that died. We analysed a subset of the dataset, including patients who had been discharged by 30 April 2021, in order to ensure at least 30 days of follow-up post discharge for all patients. We included admission to ICU as an additional explanatory variable in this analysis.

Multivariable models were obtained by forward model selection and AIC comparison. Virus variant was kept in all multivariable models regardless of statistical significance. AIC comparison was also used for determining whether age was included linearly, or with a spline. Adjusted HR (aHR) and odds-ratios (aOR) were reported. Estimates from all univariable and multivariable models are presented in supplementary materials B.

In order to check the robustness of our estimates, we conducted sensitivity analyses by changing our study population (for example, only including cases who had WGS results), and by adjusting our outcome definitions (for example, calculating LoS excluding all the time between hospital stays) to further explore if our main results were robust (supplementary materials A, part 3).

### Ethics

Ethical approval for this study was granted by Regional Committees for Medical Research Ethics - South East Norway, reference number 249509.

## RESULTS

### Patients infected with B.1.1.7 or a non-VOC

During the study period, 2,499 reported cases of COVID-19 were hospitalised not more than two days before and less than 28 days after a positive SARS-CoV-2 test. Of these, 142 (6%) were vaccinated with at least one dose of a COVID-19 vaccine before sampling or hospitalisation and excluded from the analysis. We also excluded three patients who had a reported stay in ICU outside of their hospital stay, due to assumed incomplete information on hospital stays.

Of the remaining 2,354 patients (94%), 1186 (50%) had known virus variant. Few differences were observed between patients who had known virus variant and those who did not (supplementary materials A, part 2.1). The key difference was that the proportion of cases tested for variants was higher among patients admitted to ICU than among those not admitted to ICU (57% vs 49%, p = 0.007). Patients tested for variants were also younger (median age 53 vs. 55, p = 0.005).

Of the 1,186 patients, 946 (81%) were classified as B.1.1.7 and 157 (13%) as non-VOC and formed the study cohort, while 27 (2%) were another VOC and 53 (4%) could not be classified. Characteristics of the 1,103 patients infected with B.1.1.7 or a non-VOC are presented in table 1. The proportion of B.1.1.7 in the study cohort increased throughout the study period from 0% in week 52, 2020 to 41% in week 5, 2021 and 88% in week 7, 2021. From week 11, 2021 onwards, 99% of patients were B.1.1.7. The median time from testing to hospitalisation was 4 days (IQR: 0–8) for non-VOC and 6 days (IQR: 2–8) for B.1.1.7 (p-value 0.13). At the end of the study period 16 patients (1.5%) were still admitted to hospital, all B.1.1.7. Of those 16, two had not been admitted to ICU, eight were still admitted to ICU and six had been discharged from ICU. Of 62 deaths in hospital, 27 had not been admitted to ICU, 31 died in ICU and four died in hospital after discharge from ICU.

**Table 1.**
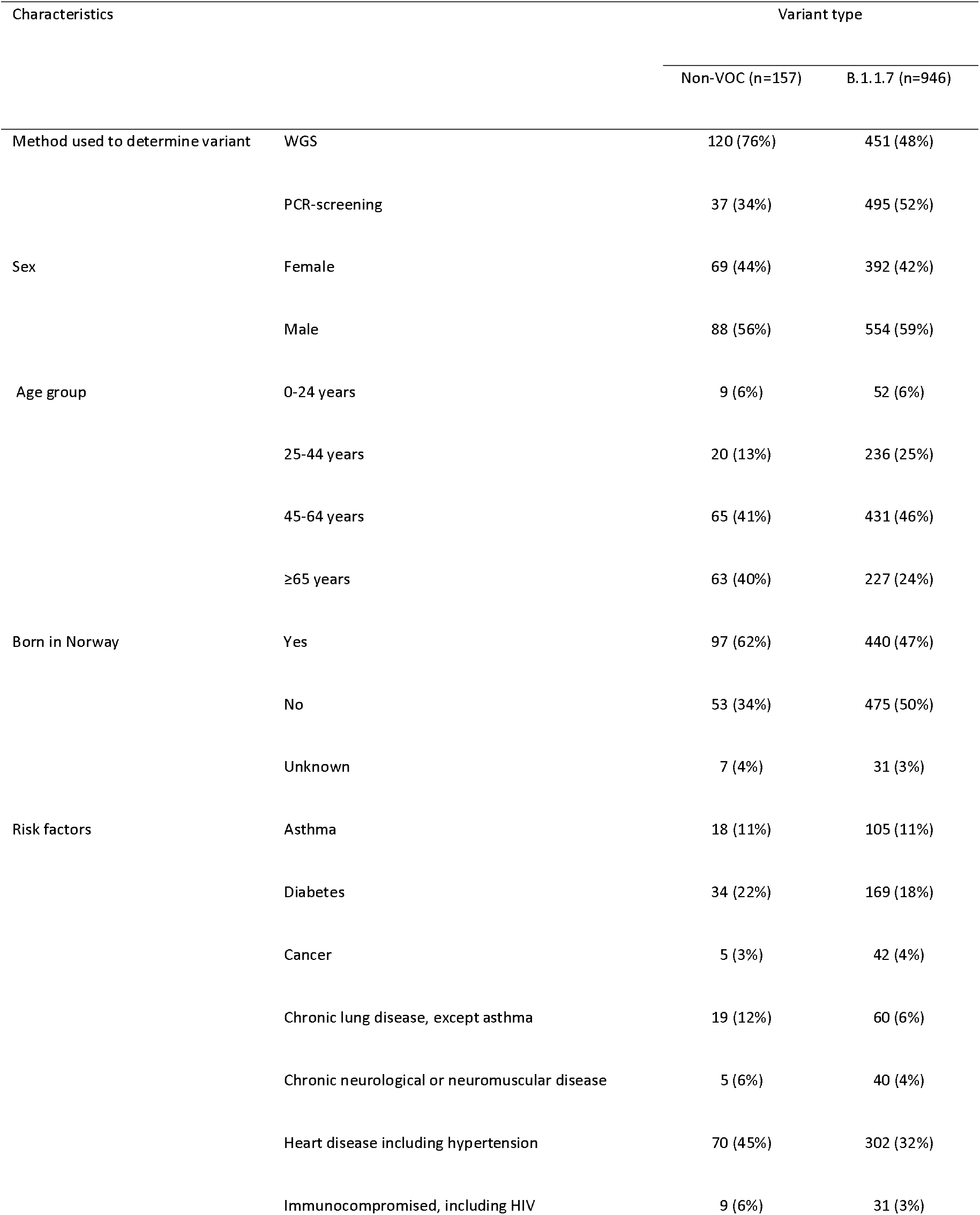
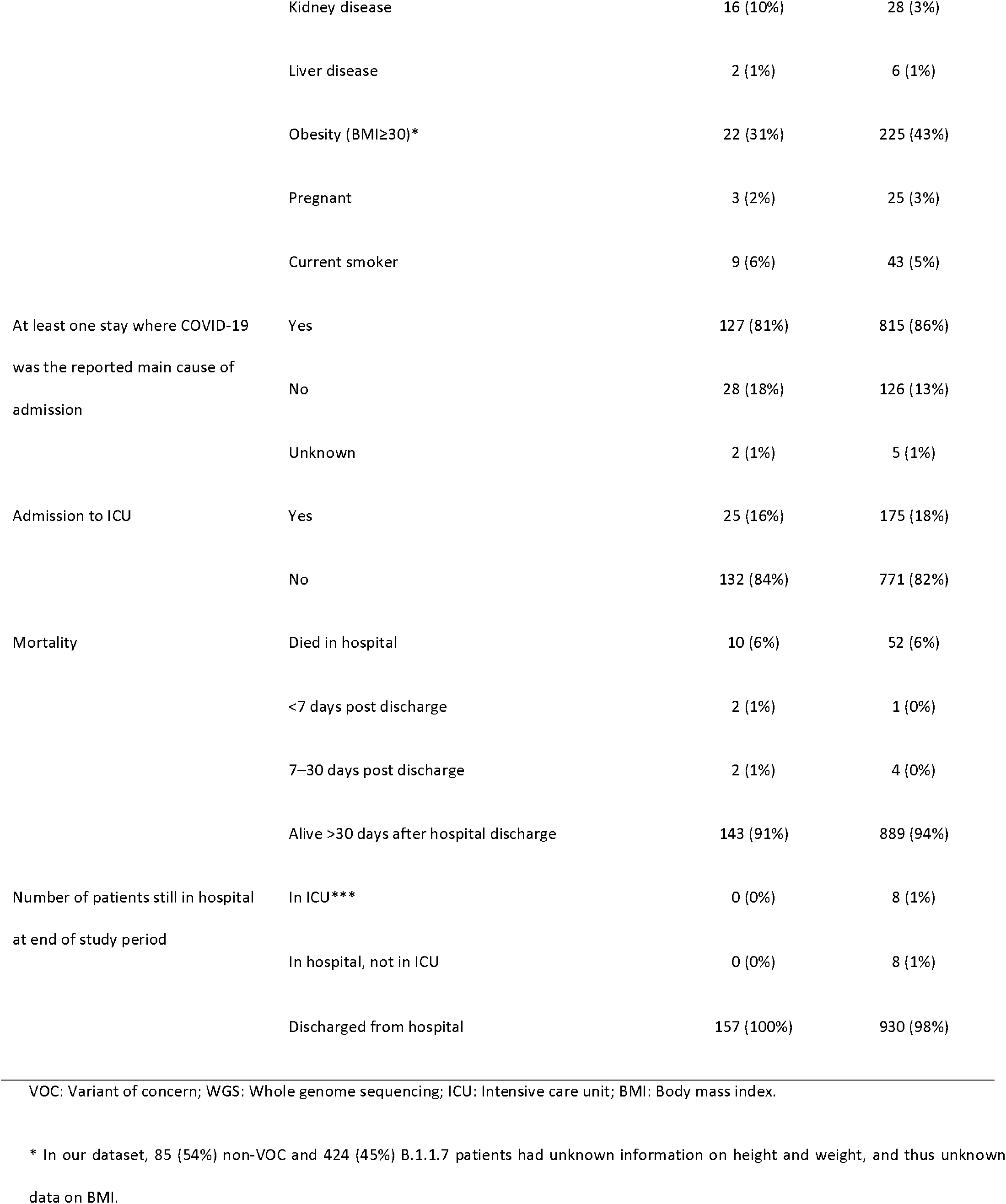
Characteristics of hospitalised SARS-CoV-2 positive patients infected with B.1.1.7 or a non-VOC, Norway, 21 December 2020 – 25 April 2021.

### Time from symptom onset to hospital admission

Date of symptom onset was known for 445 (47%) B.1.1.7 patients, and 93 (60%) non-VOC patients. Few differences were observed between patients who had known time of symptom onset and those who did not (supplementary materials A, part 2.2). The key difference was in county of residence, which is included in the final multivariable model (supplementary material B). The median time from symptom onset to hospital admission was 8 days (IQR: 5–10) for B.1.1.7 patients, and 8 days (IQR: 4– 11) for non-VOC patients (table 2). In the univariable analysis, the time from symptom onset to hospital admission was shorter for B.1.1.7, compared to non-VOC, although this was not statistically significant (HR 1.22; 95%CI 0.97–1.52), an association which was sustained in the multivariable model (aHR 1.21; 95%CI 0.94–1.55) (table 2, figure 1).

**Table 2.** Crude and adjusted hazard ratios from survival analysis for time from symptom onset to hospital admission, and length of stay in hospital and intensive care, among hospitalised SARS-CoV-2 positive patients infected with B.1.1.7 compared to a non-VOC, Norway, 21 December 2020 – 25 April 2021.

| Outcome | Variant type |  |  |  | Crude hazard ratio for<br>B.1.1.7 compared to<br>non-VOC (95%CI) | Adjusted^ hazard ratio<br>for B.1.1.7 compared<br>to non-VOC (95%CI) |
| --- | --- | --- | --- | --- | --- | --- |
|  | Non-VOC |  | B.1.1.7 |  |  |  |
|  | Number of | Median (IQR) | Number of | Median (IQR) |  |  |
|  | patients |  | patients |  |  |  |
| Days from symptom onset to<br>hospital admission* | 93 | 8 (4 – 11) | 445 | 8 (5 – 10) | 1.22 (0.97 – 1.52) | 1.21 (0.94 – 1.55) |
| Days in hospital for patients<br>not admitted to ICU | 132 | 4.1 (2.1 – 7.5) | 771 | 4.0 (2.1 – 6.8) | 1.08 (0.90 – 1.31) | 0.96 (0.79 – 1.17) |
| Days in hospital before<br>admission to ICU | 25 | 2.1 (0.1 – 4.7) | 175 | 1.2 (0.2 – 3.7) | 1.28 (0.83 – 1.96) | 1.03 (0.67 – 1.59) |
| Days in ICU | 25 | 11.0 (7.2 – 16.4) | 175 | 10.6 (5.4 – 19.6) | 0.97 (0.61 – 1.56) | 0.83 (0.51 – 1.34) |
| Days in hospital after<br>discharge from ICU** | 20 | 7.2 (3.5 – 11.3) | 141 | 5.9 (3.2 – 9.8) | 1.06 (0.65 – 1.71) | 1.00 (0.61 – 1.63) |

VOC: Variant of concern; ICU: Intensive care unit; IQR: Interquartile range; 95%CI: 95% confidence interval.
<sup>^</sup> Adjusted for age, sex, county of residence, week of admission, regional health authority, country of birth, main cause of hospitalisation and underlying risk factors. The variables included in the final multivariable model were obtained by forward model selection and AIC comparison (see supplementary material B).
\* Number of patients with known date of symptom onset: non-VOC 93/157 (60%); B.1.1.7 445/946 (47%).
\*\* Excludes eight B.1.1.7 patients who were still admitted to ICU at the end of the study period, and five non-VOC and 26 B.1.1.7 who passed away in ICU.

**Figure 1.**
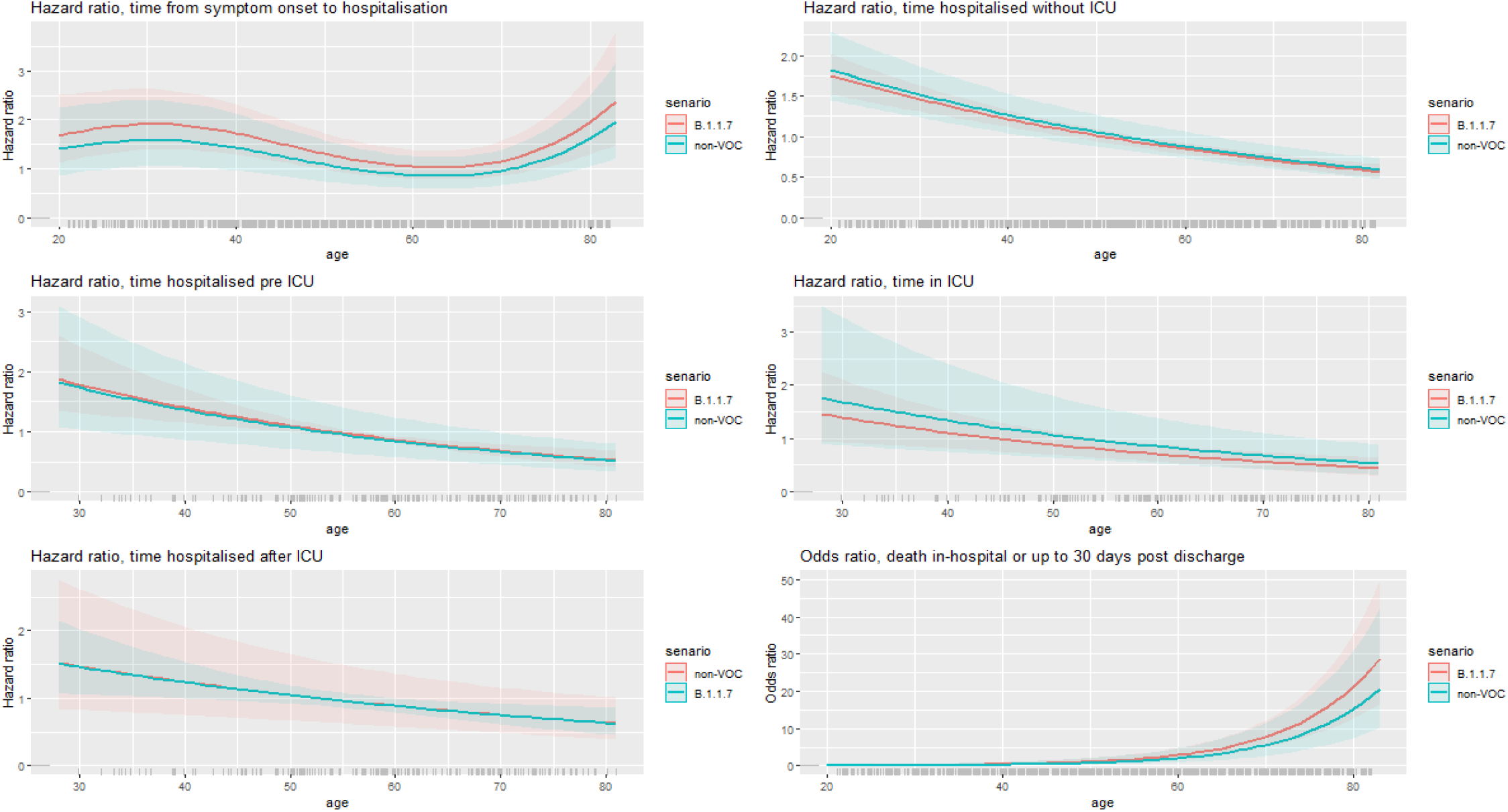
Adjusted hazard ratios (survival analysis) and adjusted odds ratios (logistic regression) for patient trajectories among hospitalised SARS-CoV-2 positive patients infected with B.1.1.7 compared to a non-VOC, Norway, 21 December 2020 – 25 April 2021. VOC: Variant of concern. To make the figure legible, not all variables controlled for in each multivariable analysis are presented. Here we present only adjusted estimates for B.1.1.7 and non-VOC. All variables included in the final multivariable models, with related hazard or odds ratios are presented in supplementary material B. In the figure, age 53 years (median age in dataset) was chosen as baseline and therefore has hazard/odds ratio equal to 1.

### Length of stay in hospital and ICU

The median overall LoS in hospital among all patients, regardless of ICU admission, was 5.0 days (IQR: 2.6–10.0) for B.1.1.7 patients and 5.1 days (IQR: 2.5–9.9) for non-VOC patients. There were minimal differences in the median LoS in hospital or ICU for B.1.1.7 patients compared to non-VOC patients (table 2). In both the univariable and multivariable models, we did not observe a difference in the LoS in hospital or ICU for B.1.1.7 patients compared to non-VOC patients (table 2, figure 1).

### Mortality

Of the 1,103 patients, 1,037 (94%) were discharged by 30 April 2021; 880 B.1.1.7 and 157 non-VOC (table 3). For B.1.1.7, 50 patients died in hospital (6%), one died less than seven days post discharge (0.1%), and three died 7–30 days post discharge (0.3%). For non-VOC, 10 patients died in hospital (6%), two died less than seven days post discharge (1.3%), and two died 7–30 days post discharge (1.3%). In both the univariable and multivariable models, we did not observe a difference in the odds of death for B.1.1.7 patients compared to non-VOC patients (table 3, figure 1).

**Table 3.** Crude and adjusted odds ratios from logistic regression for death in-hospital or up to 30 days post discharge, among hospitalised SARS-CoV-2 positive patients infected with B.1.1.7 compared to a non-VOC, 21 December 2020 – 25 April 2021, Norway.

| Outcome | Variant type |  |  |  | Crude odds ratio for<br>B.1.1.7 compared to<br>non-VOC (95%CI) | Adjusted^ odds ratio<br>for B.1.1.7 compared<br>to non-VOC (95%CI) |
| --- | --- | --- | --- | --- | --- | --- |
|  | Non-VOC |  | B.1.1.7 |  |  |  |
|  | No (%) | Yes (%) | No (%) | Yes (%) |  |  |
| Death in-hospital or up to 30 days post discharge* | 143 (91%) | 14 (9%) | 826 (94%) | 54 (6%) | 0.67 (0.36 – 1.23) | 1.39 (0.68 – 3.01) |

VOC: Variant of concern; 95%CI: 95% confidence interval.
<sup>^</sup> Adjusted for age, sex, county of residence, week of admission, regional health authority, country of birth, main cause of hospitalisation underlying risk factors and admission to ICU. The variables included in the final multivariable model were obtained by forward model selection and AIC comparison (see supplementary material B).
\* Death in-hospital or up to 30 days post discharge is limited to patients who had been discharged by 30 April 2021 (157 non-VOC, 880 B.1.1.7), in order to ensure at least 30 days of follow-up post discharge for all patients.

## DISCUSSION

In this study, we have analysed individual-level data on an unvaccinated cohort of 1,103 hospitalised COVID-19 patients in Norway, as well as demographic characteristics and underlying risk factors. Although elective surgeries in some regions were postponed during a surge in hospitalisations among COVID-19 cases in mid-March, hospitals in Norway functioned within capacity during the study period, while there were no major changes in treatment guidelines for SARS-CoV-2 patients in hospital or ICU.

Our findings indicate that there is no difference in the LoS in hospital and ICU, and odds of mortality up to 30 days post discharge for persons infected with B.1.1.7 compared to non-VOC. These findings are in line with other published studies. A study based on a cohort of 341 hospitalised patients in London, UK, found no evidence of an association between B.1.1.7 and severe disease or death (14). Further studies from the UK based on cohorts ranging from 80 to 1,310 patients have found similar results (15-17), while a study based on 3,432 ICU patients did not find increased acute severity of illness on admission to ICU, duration of organ support, duration of critical care or mortality among patients infected with B.1.1.7 (8). This suggests that, while B.1.1.7 seems to increase the risk of hospitalisation (3, 6, 9-11), other patient characteristics, such as age and underlying risk factors, determine patient trajectories and healthcare required among those hospitalised with COVID-19. COVID-19 has been shown to have a clinical course consisting of several phases. Critical disease and subsequent respiratory failure are seen primarily in a later inflammatory phase, while the initial viral phase typically causes a milder clinical picture (18). B.1.1.7 has been shown to cause a more rapid replication in the initial viral replication phase than the original Wuhan wild-type virus (19) and patients admitted to hospital with B.1.1.7 have been reported to have higher viral loads (14). One might speculate that these factors lead to increased hospital admission due to B.1.1.7 in the initial viral replication phase, but it is unclear whether the rate of development of an inflammatory phase, causing critical disease and presumably ICU admissions and mortality, is increased compared to non-VOC. We did not have access to necessary variables to explore clinical differences between patients infected with different variants of SARS-CoV-2. To further explore the observed associations, there is a clear need for further studies, particularly on larger patient cohorts and from a variety of settings considering local epidemiological characteristics.

The aforementioned study from London reported no difference in time from symptom onset to hospitalisation for B.1.1.7 compared to other variants (14). In our study, the time from symptom onset to hospitalisation was marginally shorter for B.1.1.7 patients, a finding which was on the cusp of statistical significance in both univariable and multivariable models. Seventeen patients had a date of onset on their date of admission. Among them, there could be some nosocomial SARS-CoV-2 cases, which could underestimate this parameter, however, we consider the potential bias introduced to be minimal.

The COVID-19 pandemic has put unprecedented strain on health systems, and the VOC B.1.1.7 is currently the predominant circulating strain in many countries around the world. While ongoing vaccination programmes have proved effective in reducing the incidence of infection and severe disease from both VOC and non-VOC strains, the emergence of new variants will remain an area of substantial concern as we continue to battle the spread of SARS-CoV-2. The VOC B.1.617.2 (delta variant) has been reported to be more transmissible than B.1.1.7, and has already outcompeted B.1.1.7 in some countries (20, 21). B.1.617.2 has also been associated with increased risk of hospitalisation compared to B.1.1.7 in England and Scotland, although vaccines have been reported to be effective in reducing the risk of B.1.617.2 infection and hospitalisation (20-22). In Norway, spread of B.1.617.2 is currently low, but increasing, while the vaccination programme is progressing, with vaccination coverage steadily increasing, and all adults 18 years and over expected to be offered their first dose by August 2021 (2).

Norway is a country with both high testing activity and sequencing capacity. Almost half of all patients in the study period had known data on variant, and our results were robust when we conducted various sensitivity analyses, such as restricting our study population to only cases sequenced with WGS. We also had minimal censoring of the study cohort, with 1.5% of patients still admitted to hospital at the end of the follow-up period. Patients with known variant were slightly younger and had a fractionally higher proportion of ICU admissions than all SARS-CoV-2 positive patients in the study period. Both age and ICU admission are associated with our outcomes (23, 24), however age is included as an explanatory variable in our models, while we modelled LoS separately for patients admitted to ICU, and controlled for ICU admission in our model for mortality, thus we consider our results to be generalisable. For some analyses, the number of patients was low, which needs to be considered when interpreting our results. Finally, in our dataset, some reported risk factors do not distinguish between well-regulated or treated conditions and unregulated or untreated conditions, while 46% of patients had unknown body mass index. It is therefore likely that our model does not fully adjust for certain risk factors.

Current evidence suggests that, while B.1.1.7 seems to increase the risk of hospitalisation compared to non-VOC, it does not appear to influence hospitalised patient trajectories. These findings, along with the success of vaccination programmes, are encouraging for ongoing capacity planning in the hospital sector, particularly as societies ease lockdowns. Timely analysis on the association between current and future VOC and the risk of severe disease and impact on patient trajectories will be essential to ensure health systems remain prepared and able to appropriately respond to this evolving public health threat. These analyses need to come from a variety of settings, considering local epidemiological characteristics.

## Data Availability

The datasets analysed during the current study come from the national emergency preparedness registry for COVID-19, housed at the Norwegian Institute of Public Health. The preparedness registry is temporary and comprises data from a variety of central health registries, national clinical registries and other national administrative registries. Further information on the preparedness registry, including access to data from each individual data source, is available at https://www.fhi.no/en/id/infectious-diseases/coronavirus/emergency-preparedness-register-for-covid-19/.

## NOTES

### Authors’ contributions

RW and EAB conceived the idea for the study. RW drafted the study protocol and coordinated the study. MLS, NA, UN, KB, OH, RK and EAB contributed directly to the acquisition of data. RW, ABK, ES, BVS, LaVe, MLS, HB, NA, UN, KB and OH contributed to data cleaning, validation and preparation. RW and ABK led the data analysis. All co-authors contributed to the interpretation of the results. RW and ABK drafted the manuscript. All co-authors contributed to the revision of the manuscript and approved the final version for submission.

## Acknowledgements

First and foremost, we wish to thank all those who have helped report data to the national emergency preparedness registry at the Norwegian Institute of Public Health (NIPH) throughout the pandemic. We also highly acknowledge the efforts that regional laboratories have put into establishing a routine variant screening procedure or whole genome sequencing at short notice and registration of all analysis in national registries for surveillance. Thanks also to the staff at the Virology and Bacteriology departments at NIPH involved in national variant identification and whole genome analysis of SARS-CoV-2 viruses. We also highly acknowledge the efforts of staff at hospitals around Norway to ensure the reporting of timely and complete data to the Norwegian Intensive Care and Pandemic Registry, as well as colleagues at the register itself. We would also like to thank Anja Elsrud Schou Lindman, project director for the national preparedness registry, and all those who have enabled data transfer to this registry, especially Gutorm Høgåsen at the NIPH, who has been in charge of the establishment and administration of the registry. We would like to acknowledge Jacob Berild and Camilla Mauroy, who coordinate the surveillance of COVID-19 related deaths at the NIPH. We would like to thank Trude Marie Lyngstad, Anders Skyrud Danielsen, Nora Dotterud and Evy Dvergsdal at the NIPH for their assistance in cleaning the data from different registries.

