## Supplementary materials A for "Trajectories of hospitalisation for patients infected with SARS-CoV-2 variant B.1.1.7 in Norway, December 2020 – April 2021"

### 1. Additional information on the data sources

#### 1.1 Norwegian Intensive Care and Pandemic Registry

The Norwegian Intensive Care and Pandemic Registry (NIPaR) is a national clinical registry that was expanded to include COVID-19 patients in conjunction with the COVID-19 pandemic. In the registry, all patients who have tested positive for SARS-CoV-2 and are admitted to hospital are registered. For patients who contracted SARS-CoV-2 while admitted to hospital, the time of admission is set to the date of symptom onset, or date of sampling if the patient is asymptomatic. The date of discharge for these patients is registered as when they recovered from COVID-19, even if they are still admitted to hospital for other causes. Patients who are readmitted to hospital without symptoms are only registered if they test positive for SARS-CoV-2 again and are isolated, or if a new test is not taken and the patient is isolated. Patients who are readmitted to hospital are not registered if they have tested negative for SARS-CoV-2, test positive but do not require isolation, or a new test is not taken, and the patient is not isolated. In addition, patients who are admitted to hospital for long-term complications of COVID-19 are registered if less than three months has passed since positive test.

The reported main cause of hospitalisation is a clinical assessment. For patients reported with a different main cause than COVID-19, we cannot rule out that COVID-19 may have been a contributing factor for admission.

In NIPaR, potential risk factors diagnosed before admission are registered. The following risk factors are registered; asthma, diabetes (type 1 and 2), pregnancy, heart disease including hypertension, cancer, chronic lung disease, chronic neurological or neuromuscular disease, liver disease, immunocompromised including HIV and treatment, kidney disease, current smoker and body mass index (BMI) (calculated as weight in kilograms divided by height in centimetres squared). For cancer, only active cancer is registered, meaning cancer where the patient still receives treatment, or regular control. Other well-regulated or treated conditions are not distinguished from unregulated or untreated conditions, for example asthma. In our dataset, 509 (46%) patients had unknown information on height and weight, and thus unknown data on BMI and obesity. In our models, obesity was therefore included as a three-level categorical variable, yes, no and unknown.

Full details on the registration of hospitalised patients are available here (in Norwegian): <https://helse-bergen.no/norsk-pandemiregister/registrering-i-norsk-pandemiregister-informasjon-til-ansatte>

NIPaR also includes data on patients who have tested positive for SARS-CoV-2 and are admitted to an intensive care unit (ICU). Patients are registered as ICU patients if they fulfil one of five categories:

1. Length of stay over 24 hours in intensive care
2. Require ventilatory support
3. Are transferred between intensive care wards
4. Persistent administration of vasoactive medication
5. Length of stay under 24 hours, but passed away dur during stay in intensive care

Full details on the registration of intensive care patients are available here (in Norwegian): <https://helse-bergen.no/norsk-intensivregister-nir/korona-pa-intensiv/hvordan-registrere-covid-19>

#### 1.2 Surveillance of COVID-19 related deaths in Norway

In Norway, COVID-19 related deaths are defined as deaths among COVID-19 cases notified to the Norwegian Institute of Public Health by a physician, deaths where COVID-19 is reported as the underlying cause of death (through linkage to the Cause of Death Registry), or deaths within 30 days of sampling (through linkage to the Norwegian Population Registry). Deaths notified by a physician or within 30 days of sampling date are verified against the reported underlying cause of death in the Cause of Death Registry. Deaths due to COVID-19 are not distinguished from deaths with COVID-19. More details are available here (in Norwegian): <https://www.fhi.no/sv/smittsomme-sykdommer/corona/dags--og-ukerapporter/sporsmal-og-svar-om-koronaovervaking-og-statistikk/>.

In our dataset, all deaths among confirmed cases that occurred in-hospital or within 30 days of discharge had been classified as official COVID-19 related deaths. In addition, four patients died more than 30 days after discharge, none of which were classified as a COVID-19 related death. All patients reported to have died in-hospital were registered in both the Norwegian Pandemic Registry, and as an official COVID-19 related death by the Norwegian Institute of Public Health.

### 2. Assessment of representativeness of study population

#### 2.1 Study cohort compared to notified cases

We assessed the representativeness of our study population by comparing the characteristics of the cases with known virus variant data in our study period and notified cases. We found differences between patients with virus variant data with regards to county of residence, regional health authority, week of admission to hospital, age, the proportion with COVID-19 as the main cause of admission, and proportion admitted to ICU (table S1). We did not observe any difference in the time from symptom onset to hospitalisation, or length of stay (LoS) in hospital or ICU between cases with and without known virus variant data, although the time in hospital after discharge from ICU was slightly longer for cases with variant data (hazard ratio: 1.31; 95%CI: 1.03 – 2.20) (table S2).

Differences in county, regional health authority and week of admission to hospital reflect the evolution of the outbreak as well as the introduction of PCR screening methodology for virus variants at primary diagnostic laboratories. The proportion of patients with COVID-19 as the main cause of admission was higher among cases with known virus variant data (86% vs 84%), however, this was not a significant predictor of our outcomes in our multivariable models. Also, in our sensitivity analysis only including admissions where COVID-19 was the reported main cause, our estimates were robust (see part 3). There was a slightly higher proportion of patients aged 45-64 years among cases with known virus variant data (46% vs 43%) and slightly lower proportion among patients ≥65 years (26% vs 28%). The median age among cases with known virus variant data was 53 (IQR: 43 – 65), compared to 55 (IQR: 43 – 69) among cases without known virus variant data (Wilcoxon rank-sum test p value = 0.005). Age is included as an explanatory variable in our models. The proportion of cases admitted to ICU among cases with known virus variant data was 18%, compared to 16% among all notified cases. We modelled LoS separately for patients admitted to ICU, and controlled for ICU admission in our model for mortality.

*Table S1: Characteristics of patients with known virus variant data compared to all notified cases, Norway, 21 December 2020 – 25 April 2021.*

| **Characteristics** | | **All notified cases (%)** | **Known virus variant data** | | | |
| --- | --- | --- | --- | --- | --- | --- |
|  |  |  | **N** | | **% of all notified** | |
| Total | | 2,354 | 1186 | | 50% | |
| Sex | Female | 1,391 (59%) | 696 | | 59% | |
|  | Male | 963 (41%) | 490 | | 41% | |
|  |  |  | Chi^2^ p = 0.69 | | | |
| Age group | 0-24 years | 122 (5%) | 63 | | 5% | |
|  | 25-44 years | 543 (23%) | 278 | | 23% | |
|  | 45-64 years | 1,012 (43%) | 540 | | 46% | |
|  | ≥65 years | 677 (28%) | 305 | | 26% | |
|  |  |  | Chi^2^ p =0.009 | | | |
| Born in Norway | Yes | 1,131 (48%) | 572 | | 48% | |
|  | No | 1,123 (48%) | 570 | | 48% | |
|  | Unknown | 100 (4%) | 44 | | 4% | |
|  |  |  | Chi^2^ p = 0.43 | | | |
| Number of risk factors* | 0 | 711 (31%) | 362 | | 31% | |
|  | 1 | 838 (36%) | 430 | | 37% | |
|  | ≥2 | 781 (34%) | 381 | | 32% | |
|  |  |  | Chi^2^ p = 0.56 | | | |
| Week of admission to hospital | Week 52 | 99 (4%) | 12 | | 1% | |
|  | Week 53 | 118 (5%) | 14 | | 1% | |
|  | Week 1 | 117 (5%) | 9 | | 0.8% | |
|  | Week 2 | 109 (5%) | 21 | | 2% | |
|  | Week 3 | 101 (4%) | 22 | | 2% | |
|  | Week 4 | 57 (2%) | 20 | | 2% | |
|  | Week 5 | 50 (2%) | 22 | | 2% | |
|  | Week 6 | 56 (2%) | 34 | | 3% | |
|  | Week 7 | 66 (3%) | 45 | | 4% | |
|  | Week 8 | 85 (3%) | 70 | | 6% | |
|  | Week 9 | 116 (5%) | 98 | | 8% | |
|  | Week 10 | 198 (8%) | 147 | | 12% | |
|  | Week 11 | 246 (10%) | 138 | | 12% | |
|  | Week 12 | 215 (9%) | 112 | | 9% | |
|  | Week 13 | 235 (10%) | 130 | | 11% | |
|  | Week 14 | 194 (8%) | 105 | | 9% | |
|  | Week 15 | 165 (7%) | 99 | | 8% | |
|  | Week 16 | 127 (5%) | 88 | | 7% | |
|  |  |  | Chi^2^ p = <0.0001 | | | |
| County of residence | Agder | 48 (2%) | 29 | | 2% | |
|  | Innlandet | 86 (4%) | 60 | | 5% | |
|  | Møre and Romsdal | 44 (2%) | 19 | | 2% | |
|  | Nordland | 28 (1%) | 16 | | 1% | |
|  | Oslo | 773 (32%) | 356 | | 30% | |
|  | Rogaland | 153 (7%) | 80 | | 7% | |
|  | Troms and Finnmark | 17 (0.7%) | 6 | | 0.5% | |
|  | Trøndelag | 52 (2%) | 14 | | 1% | |
|  | Vestfold and Telemark | 143 (6%) | 71 | | 6% | |
|  | Vestland | 109 (5%) | 72 | | 6% | |
|  | Viken | 871 (37%) | 448 | | 38% | |
|  | Unknown | 30 (1%) | 15 | | 1% | |
|  |  |  |  | | Chi^2^ p = <0.0001 | |
| Regional health authority | Mid-Norway | 96 (4%) | 33 | | 3% | |
|  | North | 46 (2%) | 25 | | 2% | |
|  | South-East | 1,938 (82%) | 968 | | 82% | |
|  | West | 274 (12%) | 160 | | 13% | |
|  |  |  |  | | Chi^2^ p = 0.0001 | |
| COVID-19 the main cause of admission | Yes | 1,971 (84%) | | 1,019 | | 86% |
|  | No | 374 (16%) | | 160 | | 13% |
|  | Unknown | 9 (0.4%) | | 7 | | 0.6% |
|  |  |  | Chi^2^ p = 0.002 | | | |
| Admitted to ICU | Yes | 369 (16%) | 211 | | 18% | |
|  | No | 1,985 (84%) | 975 | | 82% | |
|  |  |  | Chi^2^ p = 0.007 | | | |
| Mortality** | Number who died in hospital | 123 (5%) | | 63 | | 6% |
|  | Number who died <7 days post discharge | 10 (0.4%) | | 3 | | 0.3% |
|  | Number who died 7 – 30 days post-discharge | 8 (0.4%) | | 5 | | 0.5% |
|  | Number alive >30 days after hospital admission | 2,108 (94%) | | 1,044 | | 94% |
|  |  |  | Chi^2^ p = 0.531 | | | |

ICU: Intensive care unit.

* Asthma, diabetes, pregnancy, heart disease including hypertension, cancer, chronic lung disease, chronic neurological or neuromuscular disease, liver disease, immunocompromised, kidney disease, smoker and obesity (BMI≥30). We also compared cases with known virus variant data and notified cases for each individual risk factor. Liver disease (all notified cases 1.3%; cases with known virus variant data 0.8%; p value = 0.03) and kidney disease (all notified cases 5%; cases with known virus variant data 4%; p value = 0.03) were more frequent among all notified cases than cases with known virus variant data.

** Only includes patients who were discharged by 30 April 2021 (notified cases n=2,249; cases with known virus variant data n=1,115).

*Table S2: Hazard ratio from survival analysis of time from symptom onset to admission to hospital, and length of stay in hospital and intensive care, among patients with known virus variant compared to cases with unknown virus variant, Norway, 21 December 2020 – 25 April 2021.*

| **Outcome** | | **Unknown virus variant** | **Known virus variant** | | |
| --- | --- | --- | --- | --- | --- |
|  |  | **Number of patients** | **Number of patients** | **Hazard ratio compared to unknown virus variant**  **(95% confidence interval)** | **P-value** |
| Days from symptom onset to admission to hospital | | 578 | 538 | 0.95 (0.84 – 1.07) | 0.37 |
| Length of stay | Hospital for patients not admitted to ICU | 1,010 | 975 | 0.98 (0.90 – 1.07) | 0.65 |
|  | Hospital before admission to ICU | 158 | 211 | 1.07 (0.87 – 1.32) | 0.51 |
|  | ICU | 158 | 211 | 0.91 (0.72 – 1.14) | 0.41 |
|  | Hospital after discharge from ICU | 129* | 170** | 1.31 (1.03 – 2.20) | 0.03 |

* 29 patients with unknown variant data died in ICU

** 33 patients with known variant data died in ICU, and eight were still in ICU at the end of the study period.

#### 2.2 Patients with known data on symptom onset compared to all patients in the study cohort

Overall, 538/1,103 patients (49%) in the analysis dataset had data on date of symptom onset. Compared to all patients in the analysis dataset, a lower proportion of patients with data on date of symptom onset were admitted to ICU (16% vs. 18%, chi-squared p value = 0.034), although among patients with known date of symptom onset we did not observe a significant difference in the time from symptom onset to hospitalisation between patients admitted to ICU and those not admitted to ICU (Wilcoxon rank-sum test p value = 0.13), indicating that this difference should not bias our results. We also observed a significant difference in county of residence. Patients with known date of symptom onset were notably more likely to come from Viken (51% with known symptom onset vs. 25% with unknown symptom onset), Rogaland (9% vs. 5%), Vestland (9% vs. 5%) and Innlandet (7% vs. 3%). Conversely, patients with unknown date of symptom onset were more likely to come from Oslo (51% with unknown symptom onset vs. 11% with known symptom onset) (chi-squared p value <0.001). We also observed a significant difference in week of hospitalisation between patients with and without data on date of symptom onset, with 52% of patients admitted between week 52 2020 – week 7 2021 having data on symptom onset, compared to 48% between week 8 – 16 2021 (chi-squared p value <0.001). Both county of residence and week of hospitalisation may bias our estimates if there are systematic differences in the time from symptom onset to hospitalisation for these variables, for example if residents from one county were more likely to seek healthcare or be admitted earlier than residents from another county, or if the potential for a person to be admitted to hospital changed over the study period for example if hospital capacity was stretched. Week of hospitalisation was not a significant predictor of time from symptom onset to hospitalisation in our multivariable models, while we controlled for county of residence in our final multivariable model.

### 3. Sensitivity analyses

In addition to the main analysis, we also conducted a number of sensitivity analyses to further explore our results. The estimates from our sensitivity analyses are presented in table S3. The methodology used was the same as the one used in the main analysis. In our main analysis, we based patient admission and discharge dates on date of admission and discharge from hospital wards. However, for nine patients admitted to ICU, their registered time of admission to ICU was before their registered date of admission to hospital. For 6 patients, this time was <3 hours, although for one patient this difference was almost one day. In addition, one patient admitted to ICU had a registered date of discharge three hours after their registered date of discharge from hospital, while an additional eight patients with a date of discharge from hospital were missing a date of discharge from ICU. We conducted a sensitivity analysis to see if assuming that the registered data from ICU records was correct instead of data from ward records would affect our results. In our main analysis, we included time between stays in hospital/ICU if it was <24 hours or <12 hours, respectively. This was to ensure that we included time for transfers between wards and hospitals in our analysis. In total, 56/1,103 patients had separate hospital stays registered <24 hours apart, and 50/200 patients had separate ICU stays registered <12 hours apart. We conducted a sensitivity analysis where we excluded all time between hospital and ICU stays in calculating the LoS per patient. We also conducted sensitivity analyses where we limited our study population to only those admitted to hospital with COVID-19 as the main cause of admission, only those whose variant data came from whole genome sequencing results, and only those who had a LoS in hospital of at least 24 hours.

*Table S3. Sensitivity analysis for time from symptom onset to admission to hospital, length of stay in hospital and intensive care (survival analysis) and odds of death in-hospital and post discharge (logistic regression), B.1.1.7 compared to non-VOC, Norway, 21 December 2020 – 25 April 2021.*

| **Analysis** | **Non-VOC** | **B.1.1.7** | |  | | | |  | |
| --- | --- | --- | --- | --- | --- | --- | --- | --- | --- |
|  | **Number of patients** | **Number of patients** | | **Days from symptom onset to admission to hospital, aHR for B.1.1.7 compared to non-VOC (95%CI)** | **Days in hospital for patients not admitted to ICU, aHR for B.1.1.7 compared to non-VOC (95%CI)** | | **Days in ICU, aHR for B.1.1.7 compared to non-VOC (95%CI)** | | **Death in-hospital or up to 30 days post discharge, aOR for B.1.1.7 compared to non-VOC (95%CI)** |
| Main analysis (n=1,103) | 157 | 946 | | 1.21 (0.94 – 1.55) | 0.96 (0.79 – 1.17) | | 0.83 (0.51 – 1.34) | | 1.39 (0.68 – 3.01) |
| Changes in definition of length of stay | | |  | | |  | | | |
| Assuming registered data from ICU records are correct (n=1,103) | 157 | 946 | | 1.21 (0.94 – 1.55) | 0.96 (0.79 – 1.17) | | 0.91 (0.55 – 1.49) | | 1.35 (0.66 – 2.91) |
| Excluding all time between stays (n=1,103) | 157 | 946 | | 1.21 (0.94 – 1.55) | 0.96 (0.79 – 1.16) | | 0.96 (0.59 – 1.57) | | 1.39 (0.68 – 3.01) |
| Changes in study population | | |  | | |  | | | |
| Only including admissions where COVID-19 was the reported main cause (n=942) | 127 | 815 | | 1.23 (0.94 – 1.62) | 0.87 (0.70 – 1.08) | | 0.84 (0.51 – 1.) | | 1.32 (0.58 – 3.26) |
| Only including WGS cases (n=571) | 120 | 451 | | 1.42 (1.02 – 1.96) | 0.86 (0.68 – 1.09) | | 0.78 (0.44 – 1.39) | | 1.45 (0.64 – 3.49) |
| Length of stay in hospital at least 24 hours (n=1,004) | 142 | 862 | | 1.26 (0.97 – 1.65) | 1.00 (0.81 – 1.23) | | 0.83 (0.51 – 1.34) | | 1.32 (0.83 – 2.18) |

VOC: Variant of concern; ICU: Intensive care unit; aHR: adjusted hazard ratio; aOR: adjusted odds ratio; 95%CI: 95% confidence intervals; WGS: whole genome sequencing. The aHR and aOR are generated controlling for the same variables as in our main analysis (see supplementary materials B).
