## Supplementary materials B for "Trajectories of hospitalisation for patients infected with SARS-CoV-2 variant B.1.1.7 in Norway, December 2020 – April 2021"

*Table 1. Crude hazard ratios (survival analysis) for time from symptom onset to hospitalisation and length of stay in hospital, hospitalised SARS-CoV-2 positive patients, Norway, 21 December 2020 – 25 April 2021.*

| variable | PreSykehus.N | PreSykehus.tid | PreSykehus.HR | SykehusIkkeICU.N | SykehusIkkeICU.tid | SykehusIkkeICU.HR |
| --- | --- | --- | --- | --- | --- | --- |
| non-VOC | 93 | 8.488 (2.917) | Ref | 132 | 5.426 (1.931) | Ref |
| B.1.1.7 | 445 | 8.015 (10.021) | 1.215 (0.969, 1.523) | 771 | 4.913 (2.417) | 1.083 (0.898, 1.307) |
| male | 305 | 8.371 (9.042) | Ref | 513 | 5.406 (2.366) | Ref |
| female | 233 | 7.678 (6.226) | 1.156 (0.974, 1.371) | 390 | 4.447 (2.371) | 1.278 (1.117, 1.462) |
| born in Norway | 279 | 8.316 (6.651) | Ref | 442 | 5.321 (1.853) | Ref |
| born outside of Norway | 245 | 7.666 (8.426) | 1.209 (1.017, 1.437) | 431 | 4.623 (2.976) | 1.179 (1.029, 1.350) |
| unknown country of birth | 14 | 10.353 (99.000) | 0.702 (0.410, 1.201) | 30 | 6.062 (2.829) | 0.779 (0.537, 1.129) |
| 2020-52 | 10 | 7.117 (0.819) | Ref | 11 | 4.940 (3.775) | Ref |
| 2020-53 | 13 | 6.712 (2.116) | 1.221 (0.535, 2.786) | 12 | 3.952 (5.434) | 1.290 (0.557, 2.986) |
| 2021-1 | 4 | NA (NA) | NA (NA, NA) | 7 | 9.931 (2.691) | 0.478 (0.182, 1.257) |
| 2021-2 | 12 | 7.766 (54.345) | 1.008 (0.435, 2.335) | 16 | 5.719 (1.233) | 0.808 (0.366, 1.784) |
| 2021-3 | 14 | 7.886 (0.924) | 0.695 (0.308, 1.573) | 21 | 4.017 (1.128) | 1.241 (0.580, 2.652) |
| 2021-4 | 7 | 6.504 (1.276) | 0.807 (0.305, 2.137) | 15 | 6.654 (0.826) | 0.789 (0.346, 1.800) |
| 2021-5 | 11 | 8.023 (26.199) | 0.882 (0.374, 2.078) | 20 | 5.891 (3.159) | 0.847 (0.391, 1.836) |
| 2021-6 | 14 | 7.828 (6.531) | 0.963 (0.428, 2.168) | 28 | 5.259 (1.680) | 0.989 (0.477, 2.051) |
| 2021-7 | 16 | 6.813 (17.229) | 0.962 (0.432, 2.142) | 35 | 4.612 (3.479) | 1.055 (0.522, 2.133) |
| 2021-8 | 26 | 7.204 (4.461) | 1.080 (0.520, 2.241) | 55 | 4.907 (1.425) | 1.017 (0.517, 1.999) |
| 2021-9 | 37 | 7.358 (3.194) | 1.016 (0.505, 2.045) | 73 | 5.637 (2.087) | 0.949 (0.489, 1.842) |
| 2021-10 | 55 | 7.009 (5.232) | 1.202 (0.612, 2.360) | 115 | 5.460 (2.216) | 0.950 (0.497, 1.815) |
| 2021-11 | 60 | 8.923 (13.354) | 0.761 (0.389, 1.488) | 92 | 4.092 (3.123) | 1.321 (0.687, 2.540) |
| 2021-12 | 48 | 6.844 (4.428) | 1.305 (0.659, 2.582) | 78 | 4.663 (2.614) | 1.227 (0.634, 2.374) |
| 2021-13 | 61 | 9.514 (99.000) | 0.714 (0.365, 1.394) | 100 | 5.281 (2.499) | 0.949 (0.495, 1.821) |
| 2021-14 | 60 | 8.148 (21.368) | 0.937 (0.479, 1.832) | 78 | 4.733 (3.463) | 1.139 (0.589, 2.201) |
| 2021-15 | 54 | 8.555 (99.000) | 0.926 (0.471, 1.819) | 79 | 3.997 (2.493) | 1.316 (0.680, 2.546) |
| 2021-16 | 36 | 8.901 (7.202) | 0.675 (0.333, 1.368) | 68 | 4.591 (2.678) | 1.208 (0.621, 2.348) |
| 0-24 years old | 23 | 5.787 (1.319) | Ref | 57 | 2.213 (1.708) | Ref |
| 25-44 years old | 123 | 7.555 (9.289) | 0.955 (0.609, 1.498) | 229 | 3.691 (3.444) | 0.683 (0.510, 0.915) |
| 45-64 years old | 252 | 8.625 (19.512) | 0.738 (0.480, 1.135) | 395 | 5.127 (2.696) | 0.465 (0.352, 0.616) |
| 65+ years old | 140 | 7.734 (3.796) | 0.831 (0.533, 1.296) | 222 | 6.768 (2.743) | 0.328 (0.244, 0.441) |
| not known risk factor | 164 | 8.737 (9.818) | Ref | 298 | 3.836 (2.293) | Ref |
| known risk factor | 369 | 7.774 (6.859) | 1.245 (1.035, 1.497) | 594 | 5.586 (2.549) | 0.640 (0.556, 0.738) |
| unknown risk factor | 5 | 7.437 (9.144) | 1.152 (0.473, 2.807) | 11 | 3.822 (1.953) | 0.734 (0.402, 1.343) |
| not asthma | 480 | 8.108 (6.721) | Ref | 805 | 5.019 (2.257) | Ref |
| asthma | 58 | 7.868 (25.961) | 1.178 (0.896, 1.549) | 98 | 4.724 (3.103) | 1.048 (0.848, 1.294) |
| not diabetes | 439 | 8.290 (8.156) | Ref | 754 | 4.702 (2.263) | Ref |
| diabetes | 99 | 7.120 (5.198) | 1.384 (1.111, 1.725) | 149 | 6.324 (3.397) | 0.704 (0.588, 0.843) |
| not pregnancy | 222 | 7.754 (5.865) | Ref | 367 | 4.609 (2.259) | Ref |
| pregnancy | 11 | 6.336 (99.000) | 1.775 (0.966, 3.263) | 23 | 2.588 (99.000) | 2.128 (1.391, 3.256) |
| Male (related to pregnancy) | 305 | 8.371 (9.042) | 0.883 (0.743, 1.050) | 513 | 5.406 (2.366) | 0.807 (0.704, 0.926) |
| not heart disease including hypertension | 359 | 8.355 (8.236) | Ref | 624 | 4.528 (2.232) | Ref |
| heart disease including hypertension | 179 | 7.505 (6.332) | 1.241 (1.036, 1.486) | 279 | 6.014 (2.806) | 0.700 (0.605, 0.810) |
| not cancer | 519 | 8.133 (8.841) | Ref | 864 | 4.914 (2.376) | Ref |
| cancer | 19 | 5.810 (0.563) | 1.250 (0.789, 1.979) | 39 | 7.341 (1.429) | 0.558 (0.393, 0.794) |
| not chronic lung disease | 504 | 8.191 (8.516) | Ref | 843 | 4.847 (2.368) | Ref |
| chronic lung disease | 34 | 6.215 (2.433) | 1.488 (1.051, 2.108) | 60 | 7.224 (2.325) | 0.624 (0.470, 0.829) |
| not chronic neurological or neuromuscular disease | 514 | 8.158 (8.038) | Ref | 860 | 4.903 (2.361) | Ref |
| chronic neurological or neuromuscular disease | 24 | 6.225 (2.310) | 1.595 (1.058, 2.405) | 43 | 6.820 (2.087) | 0.680 (0.493, 0.939) |
| not immunocompromised | 523 | 8.128 (7.903) | Ref | 876 | 4.951 (2.329) | Ref |
| immunocompromised | 15 | 6.425 (0.818) | 1.658 (0.991, 2.772) | 27 | 6.176 (2.110) | 0.667 (0.444, 1.002) |
| not kidney disease | 519 | 8.082 (8.098) | Ref | 871 | 4.864 (2.355) | Ref |
| kidney disease | 19 | 7.980 (1.597) | 0.951 (0.601, 1.505) | 32 | 9.257 (2.586) | 0.450 (0.302, 0.670) |
| not current smoker | 520 | 8.140 (8.103) | Ref | 860 | 4.944 (2.335) | Ref |
| current smoker | 18 | 5.795 (1.510) | 1.614 (1.007, 2.585) | 43 | 5.805 (2.282) | 0.823 (0.599, 1.131) |
| not obesity | 158 | 7.791 (5.477) | Ref | 272 | 4.962 (2.451) | Ref |
| obesity | 137 | 8.493 (10.644) | 0.848 (0.673, 1.068) | 182 | 5.272 (2.701) | 0.944 (0.781, 1.142) |
| unknown obesity | 243 | 8.012 (8.135) | 0.984 (0.805, 1.203) | 449 | 4.861 (2.091) | 1.018 (0.873, 1.186) |
| Oslo | 57 | 6.183 (2.067) | Ref | 266 | 4.940 (2.073) | Ref |
| Agder | 15 | 8.944 (3.229) | 0.477 (0.269, 0.845) | 25 | 5.011 (6.406) | 0.882 (0.581, 1.340) |
| Innlandet | 38 | 8.693 (7.494) | 0.567 (0.376, 0.856) | 41 | 5.306 (2.045) | 0.923 (0.662, 1.288) |
| Møre Og Romsdal | 5 | 3.556 (1.199) | 2.801 (1.117, 7.025) | 11 | 5.030 (4.021) | 0.951 (0.520, 1.740) |
| Nordland, Troms Og Finnmark | 10 | 7.533 (22.126) | 0.800 (0.408, 1.567) | 10 | 5.255 (1.794) | 0.863 (0.458, 1.623) |
| Rogaland | 49 | 8.391 (26.908) | 0.642 (0.438, 0.941) | 65 | 4.793 (1.816) | 1.067 (0.810, 1.406) |
| Trøndelag | 8 | 8.693 (7.255) | 0.652 (0.311, 1.367) | 11 | 4.145 (2.887) | 1.143 (0.625, 2.091) |
| Vestfold Og Telemark | 35 | 7.859 (3.089) | 0.599 (0.392, 0.916) | 55 | 4.817 (3.076) | 0.995 (0.741, 1.338) |
| Vestland | 43 | 8.447 (43.297) | 0.586 (0.393, 0.875) | 51 | 2.984 (2.367) | 1.840 (1.360, 2.488) |
| Viken | 275 | 8.252 (9.660) | 0.621 (0.466, 0.827) | 361 | 5.149 (2.540) | 0.907 (0.771, 1.065) |
| Unknown county | 3 | NA (NA) | NA (NA, NA) | 7 | 6.295 (8.483) | 0.759 (0.358, 1.608) |
| South-east Health Authority | 422 | 8.032 (6.052) | Ref | 752 | 5.120 (2.428) | Ref |
| West Health Authority | 94 | 8.394 (36.915) | 0.964 (0.770, 1.208) | 119 | 4.007 (1.857) | 1.386 (1.138, 1.687) |
| Mid-Norway Health Authority | 13 | 6.945 (2.451) | 1.429 (0.822, 2.486) | 21 | 4.859 (2.891) | 1.094 (0.709, 1.689) |
| North Health Authority | 9 | 8.026 (99.000) | 1.152 (0.594, 2.234) | 11 | 4.667 (3.846) | 1.109 (0.611, 2.012) |
| COVID-19 main cause of hospitalisation | 467 | 8.441 (14.927) | Ref | 754 | 4.925 (2.764) | Ref |
| COVID-19 not main cause of hospitalisation | 67 | 5.075 (1.024) | 1.647 (1.269, 2.137) | 143 | 5.683 (1.114) | 0.833 (0.691, 1.005) |
| unknown main cause | 4 | NA (NA) | NA (NA, NA) | 6 | 2.541 (6.646) | 1.916 (0.857, 4.285) |

*Table 2. Crude hazard ratios (survival analysis) for length of stay in intensive care (ICU), hospitalised SARS-CoV-2 positive patients, Norway, 21 December 2020 – 25 April 2021.*

| variable | SykehusPreICU.N | SykehusPreICU.tid | SykehusPreICU.HR | SykehusICU.N | SykehusICU.tid | SykehusICU.HR | SykehusPostICU.N | SykehusPostICU.tid | SykehusPostICU.HR |
| --- | --- | --- | --- | --- | --- | --- | --- | --- | --- |
| non-VOC | 25 | 2.845 (1.263) | Ref | 25 | 13.627 (2.142) | Ref | 20 | 8.111 (1.907) | Ref |
| B.1.1.7 | 175 | 2.286 (1.183) | 1.275 (0.831, 1.958) | 175 | 17.010 (1.226) | 0.973 (0.609, 1.556) | 141 | 7.248 (2.234) | 1.056 (0.650, 1.713) |
| male | 129 | 2.415 (1.092) | Ref | 129 | 16.913 (1.262) | Ref | 105 | 7.497 (1.751) | Ref |
| female | 71 | 2.259 (1.661) | 1.029 (0.768, 1.378) | 71 | 16.043 (1.315) | 0.961 (0.694, 1.331) | 56 | 7.261 (3.031) | 1.015 (0.725, 1.421) |
| born in Norway | 95 | 2.991 (0.963) | Ref | 95 | 16.116 (1.325) | Ref | 76 | 6.957 (1.581) | Ref |
| born outside of Norway | 97 | 1.904 (2.294) | 1.402 (1.046, 1.880) | 97 | 17.662 (1.244) | 0.929 (0.677, 1.276) | 78 | 7.822 (2.984) | 0.860 (0.619, 1.194) |
| unknown country of birth | 8 | 1.344 (0.383) | 2.024 (0.978, 4.191) | 8 | 10.315 (0.995) | 1.064 (0.488, 2.317) | 7 | 6.049 (1.323) | 0.936 (0.429, 2.040) |
| 2020-52 | 1 | NA (NA) | NA (NA, NA) | 1 | NA (NA) | NA (NA, NA) | 1 | NA (NA) | NA (NA, NA) |
| 2020-53 | 2 | NA (NA) | NA (NA, NA) | 2 | NA (NA) | NA (NA, NA) | 0 | NA (NA) | NA (NA, NA) |
| 2021-1 | 2 | NA (NA) | NA (NA, NA) | 2 | NA (NA) | NA (NA, NA) | 2 | NA (NA) | NA (NA, NA) |
| 2021-2 | 5 | 2.623 (1.831) | 0.136 (0.016, 1.196) | 5 | 11.451 (8.227) | 0.017 (0.007, 0.040) | 5 | 6.103 (1.228) | 0.299 (0.033, 2.716) |
| 2021-3 | 1 | NA (NA) | NA (NA, NA) | 1 | NA (NA) | NA (NA, NA) | 0 | NA (NA) | NA (NA, NA) |
| 2021-4 | 4 | NA (NA) | NA (NA, NA) | 4 | NA (NA) | NA (NA, NA) | 2 | NA (NA) | NA (NA, NA) |
| 2021-5 | 2 | NA (NA) | NA (NA, NA) | 2 | NA (NA) | NA (NA, NA) | 1 | NA (NA) | NA (NA, NA) |
| 2021-6 | 4 | NA (NA) | NA (NA, NA) | 4 | NA (NA) | NA (NA, NA) | 2 | NA (NA) | NA (NA, NA) |
| 2021-7 | 8 | 1.887 (2.633) | 0.214 (0.026, 1.747) | 8 | 13.928 (5.976) | 0.015 (0.007, 0.032) | 7 | 7.056 (4.454) | 0.230 (0.027, 1.957) |
| 2021-8 | 10 | 1.979 (0.918) | 0.211 (0.026, 1.683) | 10 | 15.414 (1.451) | 0.015 (0.008, 0.029) | 9 | 6.585 (99.000) | 0.437 (0.055, 3.491) |
| 2021-9 | 16 | 2.150 (2.621) | 0.186 (0.024, 1.439) | 16 | 11.108 (5.575) | 0.020 (0.012, 0.035) | 14 | 7.191 (1.881) | 0.349 (0.045, 2.699) |
| 2021-10 | 24 | 2.316 (1.918) | 0.180 (0.024, 1.362) | 24 | 18.907 (1.336) | 0.013 (0.008, 0.021) | 21 | 9.204 (1.685) | 0.271 (0.036, 2.056) |
| 2021-11 | 27 | 2.959 (0.927) | 0.143 (0.019, 1.086) | 27 | 16.389 (1.516) | 0.014 (0.008, 0.022) | 20 | 6.332 (3.423) | 0.428 (0.057, 3.228) |
| 2021-12 | 25 | 2.626 (1.141) | 0.155 (0.020, 1.172) | 25 | 23.510 (0.596) | 0.011 (0.007, 0.019) | 18 | 6.041 (1.209) | 0.427 (0.056, 3.253) |
| 2021-13 | 22 | 1.053 (99.000) | 0.279 (0.037, 2.105) | 22 | 12.901 (1.236) | 0.021 (0.013, 0.033) | 21 | 5.842 (1.869) | 0.419 (0.055, 3.172) |
| 2021-14 | 18 | 2.916 (1.315) | 0.158 (0.021, 1.211) | 18 | 23.109 (0.668) | 0.012 (0.007, 0.021) | 14 | 10.939 (0.681) | 0.222 (0.028, 1.756) |
| 2021-15 | 14 | 0.952 (99.000) | 0.377 (0.049, 2.919) | 14 | 9.928 (4.503) | 0.024 (0.013, 0.044) | 12 | 5.610 (9.812) | 0.371 (0.048, 2.895) |
| 2021-16 | 15 | 1.660 (3.068) | 0.271 (0.035, 2.092) | 15 | 10.574 (1.123) | 0.017 (0.009, 0.030) | 12 | 5.903 (37.589) | 0.403 (0.051, 3.163) |
| 0-24 years old | 4 | NA (NA) | NA (NA, NA) | 4 | NA (NA) | NA (NA, NA) | 4 | NA (NA) | NA (NA, NA) |
| 25-44 years old | 27 | 1.450 (4.192) | 0.289 (0.101, 0.833) | 27 | 11.420 (1.469) | 0.298 (0.102, 0.867) | 24 | 6.023 (2.558) | 0.374 (0.127, 1.104) |
| 45-64 years old | 101 | 2.060 (1.555) | 0.184 (0.067, 0.507) | 101 | 15.701 (1.499) | 0.237 (0.086, 0.655) | 85 | 6.813 (2.366) | 0.381 (0.137, 1.063) |
| 65+ years old | 68 | 3.402 (1.124) | 0.134 (0.048, 0.373) | 68 | 22.451 (1.028) | 0.187 (0.067, 0.527) | 48 | 9.774 (1.875) | 0.234 (0.081, 0.671) |
| not known risk factor | 36 | 1.948 (2.439) | Ref | 36 | 17.006 (1.149) | Ref | 33 | 6.941 (2.045) | Ref |
| known risk factor | 162 | 2.474 (1.085) | 0.817 (0.567, 1.178) | 162 | 16.249 (1.328) | 0.918 (0.624, 1.350) | 126 | 7.431 (2.299) | 0.986 (0.657, 1.481) |
| unknown risk factor | 2 | NA (NA) | NA (NA, NA) | 2 | NA (NA) | NA (NA, NA) | 2 | NA (NA) | NA (NA, NA) |
| not asthma | 175 | 2.351 (1.117) | Ref | 175 | 16.257 (1.295) | Ref | 141 | 7.505 (2.018) | Ref |
| asthma | 25 | 2.413 (2.330) | 0.957 (0.628, 1.458) | 25 | 19.293 (1.179) | 0.803 (0.501, 1.285) | 20 | 6.888 (3.409) | 1.149 (0.716, 1.842) |
| not diabetes | 146 | 2.308 (1.401) | Ref | 146 | 15.990 (1.249) | Ref | 120 | 7.164 (1.930) | Ref |
| diabetes | 54 | 2.467 (0.950) | 1.028 (0.750, 1.408) | 54 | 18.368 (1.398) | 0.829 (0.581, 1.182) | 41 | 7.941 (3.318) | 0.844 (0.583, 1.221) |
| not pregnancy | 66 | 2.252 (1.517) | Ref | 66 | 17.351 (1.359) | Ref | 51 | 7.525 (2.939) | Ref |
| pregnancy | 5 | 1.992 (99.000) | 1.148 (0.460, 2.864) | 5 | 3.754 (4.519) | 5.494 (2.142, 14.089) | 5 | 4.123 (99.000) | 1.951 (0.771, 4.936) |
| Male (related to pregnancy) | 129 | 2.415 (1.092) | 0.981 (0.727, 1.323) | 129 | 16.913 (1.262) | 1.123 (0.803, 1.571) | 105 | 7.497 (1.751) | 1.032 (0.729, 1.462) |
| not heart disease including hypertension | 107 | 2.199 (1.475) | Ref | 107 | 14.714 (1.482) | Ref | 92 | 6.913 (2.297) | Ref |
| heart disease including hypertension | 93 | 2.543 (1.112) | 0.932 (0.704, 1.233) | 93 | 19.003 (1.129) | 0.845 (0.618, 1.155) | 69 | 7.996 (2.050) | 0.885 (0.639, 1.226) |
| not cancer | 192 | 2.187 (1.250) | Ref | 192 | 16.430 (1.256) | Ref | 159 | 7.386 (2.126) | Ref |
| cancer | 8 | 6.619 (7.414) | 0.281 (0.131, 0.605) | 8 | 16.380 (4.338) | 0.358 (0.089, 1.449) | 2 | NA (NA) | NA (NA, NA) |
| not chronic lung disease | 181 | 2.368 (1.226) | Ref | 181 | 16.802 (1.283) | Ref | 148 | 7.602 (2.286) | Ref |
| chronic lung disease | 19 | 2.263 (0.923) | 0.980 (0.609, 1.579) | 19 | 15.271 (1.141) | 1.022 (0.579, 1.806) | 13 | 4.681 (1.561) | 1.482 (0.819, 2.683) |
| not chronic neurological or neuromuscular disease | 194 | 2.346 (1.210) | Ref | 194 | 16.762 (1.302) | Ref | 157 | 7.360 (2.147) | Ref |
| chronic neurological or neuromuscular disease | 6 | 2.459 (0.784) | 0.927 (0.410, 2.096) | 6 | 10.304 (0.809) | 0.769 (0.283, 2.091) | 4 | NA (NA) | NA (NA, NA) |
| not immunocompromised | 187 | 2.311 (1.250) | Ref | 187 | 16.397 (1.267) | Ref | 154 | 7.296 (2.093) | Ref |
| immunocompromised | 13 | 3.729 (0.586) | 0.594 (0.327, 1.077) | 13 | 15.484 (99.000) | 0.627 (0.294, 1.339) | 7 | 9.033 (7.003) | 0.717 (0.335, 1.533) |
| not kidney disease | 188 | 2.261 (1.232) | Ref | 188 | 16.425 (1.258) | Ref | 152 | 7.320 (2.360) | Ref |
| kidney disease | 12 | 4.257 (0.883) | 0.580 (0.321, 1.047) | 12 | 19.769 (1.914) | 0.814 (0.415, 1.597) | 9 | 9.942 (0.523) | 0.777 (0.377, 1.599) |
| not current smoker | 191 | 2.301 (1.199) | Ref | 191 | 16.446 (1.278) | Ref | 156 | 7.585 (2.077) | Ref |
| current smoker | 9 | 3.659 (0.959) | 0.645 (0.329, 1.264) | 9 | 22.359 (1.233) | 0.595 (0.244, 1.454) | 5 | 4.259 (12.978) | 2.000 (0.812, 4.924) |
| not obesity | 75 | 2.280 (1.317) | Ref | 75 | 15.390 (1.522) | Ref | 59 | 8.823 (1.381) | Ref |
| obesity | 65 | 2.230 (1.242) | 1.087 (0.775, 1.525) | 65 | 16.408 (1.377) | 0.943 (0.652, 1.366) | 54 | 6.337 (2.043) | 1.455 (0.988, 2.144) |
| unknown obesity | 60 | 2.504 (1.166) | 0.998 (0.707, 1.408) | 60 | 18.149 (0.976) | 0.923 (0.630, 1.353) | 48 | 7.348 (3.554) | 1.244 (0.832, 1.860) |
| Oslo | NA | NA | NA | NA | NA | NA | NA | NA | NA |
| Agder | NA | NA | NA | NA | NA | NA | NA | NA | NA |
| Innlandet | NA | NA | NA | NA | NA | NA | NA | NA | NA |
| Møre Og Romsdal | NA | NA | NA | NA | NA | NA | NA | NA | NA |
| Nordland, Troms Og Finnmark | NA | NA | NA | NA | NA | NA | NA | NA | NA |
| Rogaland | NA | NA | NA | NA | NA | NA | NA | NA | NA |
| Trøndelag | NA | NA | NA | NA | NA | NA | NA | NA | NA |
| Vestfold Og Telemark | NA | NA | NA | NA | NA | NA | NA | NA | NA |
| Vestland | NA | NA | NA | NA | NA | NA | NA | NA | NA |
| Viken | NA | NA | NA | NA | NA | NA | NA | NA | NA |
| Unknown county | NA | NA | NA | NA | NA | NA | NA | NA | NA |
| South-east Health Authority | 160 | 2.322 (1.300) | Ref | 160 | 16.570 (1.344) | Ref | 129 | 7.047 (2.899) | Ref |
| West Health Authority | 31 | 2.286 (1.245) | 1.159 (0.786, 1.709) | 31 | 15.325 (1.297) | 1.064 (0.692, 1.635) | 25 | 12.213 (0.608) | 0.554 (0.338, 0.909) |
| Mid-Norway Health Authority | 8 | 2.721 (1.240) | 0.815 (0.399, 1.665) | 8 | 7.314 (5.134) | 1.328 (0.619, 2.850) | 7 | 5.928 (10.813) | 0.810 (0.356, 1.840) |
| North Health Authority | 1 | NA (NA) | NA (NA, NA) | 1 | NA (NA) | NA (NA, NA) | 0 | NA (NA) | NA (NA, NA) |
| COVID-19 main cause of hospitalisation | 188 | 2.328 (1.233) | Ref | 188 | 17.048 (1.342) | Ref | 152 | 7.504 (2.045) | Ref |
| COVID-19 not main cause of hospitalisation | 11 | 1.975 (1.334) | 0.747 (0.403, 1.386) | 11 | 6.379 (0.935) | 1.975 (0.965, 4.041) | 8 | 6.099 (5.709) | 0.986 (0.461, 2.111) |
| unknown main cause | 1 | NA (NA) | NA (NA, NA) | 1 | NA (NA) | NA (NA, NA) | 1 | NA (NA) | NA (NA, NA) |

*Table 3. Crude odds ratios (logistic regression) for admission to ICU and death in-hospital or up to 30 days post discharge, hospitalised SARS-CoV-2 positive patients, Norway, 21 December 2020 – 25 April 2021.*

| variable | ICU.prob (n, N) | ICU.OR | Death.prob (n, N) | Death.OR |
| --- | --- | --- | --- | --- |
| non-VOC | 0.159 (25, 157) | Ref | 0.089 (14, 157) | Ref |
| B.1.1.7 | 0.185 (175, 946) | 1.198 (0.758, 1.895) | 0.061 (54, 880) | 0.668 (0.361, 1.234) |
| male | 0.201 (129, 642) | Ref | 0.071 (43, 607) | Ref |
| female | 0.154 (71, 461) | 0.724 (0.527, 0.995) | 0.058 (25, 430) | 0.810 (0.487, 1.347) |
| born in Norway | 0.177 (95, 537) | Ref | 0.090 (46, 510) | Ref |
| born outside of Norway | 0.184 (97, 528) | 1.047 (0.766, 1.431) | 0.045 (22, 491) | 0.473 (0.280, 0.799) |
| unknown country of birth | 0.211 (8, 38) | 1.241 (0.551, 2.791) | 0.000 (0, 36) | 0.000 (0.000, Inf.000) |
| 2020-52 | 0.083 (1, 12) | Ref | 0.250 (3, 12) | Ref |
| 2020-53 | 0.143 (2, 14) | 1.833 (0.145, 23.155) | 0.143 (2, 14) | 0.500 (0.069, 3.647) |
| 2021-1 | 0.222 (2, 9) | 3.143 (0.238, 41.509) | 0.000 (0, 9) | 0.000 (0.000, Inf.000) |
| 2021-2 | 0.238 (5, 21) | 3.437 (0.352, 33.614) | 0.095 (2, 21) | 0.316 (0.045, 2.235) |
| 2021-3 | 0.045 (1, 22) | 0.524 (0.030, 9.202) | 0.091 (2, 22) | 0.300 (0.042, 2.118) |
| 2021-4 | 0.211 (4, 19) | 2.933 (0.287, 30.009) | 0.211 (4, 19) | 0.800 (0.145, 4.423) |
| 2021-5 | 0.091 (2, 22) | 1.100 (0.089, 13.545) | 0.136 (3, 22) | 0.474 (0.079, 2.826) |
| 2021-6 | 0.125 (4, 32) | 1.571 (0.158, 15.668) | 0.125 (4, 32) | 0.429 (0.080, 2.288) |
| 2021-7 | 0.186 (8, 43) | 2.514 (0.282, 22.387) | 0.023 (1, 43) | 0.071 (0.007, 0.768) |
| 2021-8 | 0.154 (10, 65) | 2.000 (0.232, 17.259) | 0.031 (2, 65) | 0.095 (0.014, 0.650) |
| 2021-9 | 0.180 (16, 89) | 2.411 (0.290, 20.035) | 0.090 (8, 89) | 0.296 (0.066, 1.321) |
| 2021-10 | 0.173 (24, 139) | 2.296 (0.283, 18.632) | 0.044 (6, 135) | 0.140 (0.030, 0.652) |
| 2021-11 | 0.227 (27, 119) | 3.228 (0.399, 26.143) | 0.070 (8, 115) | 0.224 (0.050, 0.996) |
| 2021-12 | 0.243 (25, 103) | 3.526 (0.433, 28.680) | 0.092 (9, 98) | 0.303 (0.069, 1.327) |
| 2021-13 | 0.180 (22, 122) | 2.420 (0.297, 19.733) | 0.061 (7, 115) | 0.194 (0.043, 0.884) |
| 2021-14 | 0.187 (18, 96) | 2.538 (0.308, 20.945) | 0.034 (3, 87) | 0.107 (0.019, 0.611) |
| 2021-15 | 0.151 (14, 93) | 1.949 (0.233, 16.315) | 0.037 (3, 81) | 0.115 (0.020, 0.659) |
| 2021-16 | 0.181 (15, 83) | 2.426 (0.291, 20.258) | 0.017 (1, 58) | 0.053 (0.005, 0.563) |
| 0-24 years old | 0.066 (4, 61) | Ref | 0.000 (0, 61) | Ref |
| 25-44 years old | 0.105 (27, 256) | 1.680 (0.565, 4.994) | 0.008 (2, 251) | 928905.925 (0.000, Inf.000) |
| 45-64 years old | 0.204 (101, 496) | 3.644 (1.292, 10.279) | 0.040 (18, 451) | 4807570.851 (0.000, Inf.000) |
| 65+ years old | 0.234 (68, 290) | 4.365 (1.528, 12.468) | 0.175 (48, 274) | 24562574.380 (0.000, Inf.000) |
| not known risk factor | 0.108 (36, 334) | Ref | 0.009 (3, 317) | Ref |
| known risk factor | 0.214 (162, 756) | 2.258 (1.533, 3.324) | 0.092 (65, 707) | 10.597 (3.305, 33.983) |
| unknown risk factor | 0.154 (2, 13) | 1.505 (0.321, 7.062) | 0.000 (0, 13) | 0.000 (0.000, Inf.000) |
| not asthma | 0.179 (175, 980) | Ref | 0.066 (61, 919) | Ref |
| asthma | 0.203 (25, 123) | 1.173 (0.734, 1.875) | 0.059 (7, 118) | 0.887 (0.396, 1.987) |
| not diabetes | 0.162 (146, 900) | Ref | 0.059 (50, 851) | Ref |
| diabetes | 0.266 (54, 203) | 1.872 (1.308, 2.678) | 0.097 (18, 186) | 1.716 (0.977, 3.016) |
| not pregnancy | 0.152 (66, 433) | Ref | 0.062 (25, 402) | Ref |
| pregnancy | 0.179 (5, 28) | 1.209 (0.444, 3.293) | 0.000 (0, 28) | 0.000 (0.000, Inf.000) |
| Male (related to pregnancy) | 0.201 (129, 642) | 1.398 (1.010, 1.936) | 0.071 (43, 607) | 1.150 (0.690, 1.914) |
| not heart disease including hypertension | 0.146 (107, 731) | Ref | 0.032 (22, 691) | Ref |
| heart disease including hypertension | 0.250 (93, 372) | 1.944 (1.423, 2.655) | 0.133 (46, 346) | 4.663 (2.755, 7.890) |
| not cancer | 0.182 (192, 1056) | Ref | 0.056 (56, 993) | Ref |
| cancer | 0.170 (8, 47) | 0.923 (0.425, 2.007) | 0.273 (12, 44) | 6.275 (3.066, 12.841) |
| not chronic lung disease | 0.177 (181, 1024) | Ref | 0.053 (51, 961) | Ref |
| chronic lung disease | 0.241 (19, 79) | 1.475 (0.859, 2.532) | 0.224 (17, 76) | 5.141 (2.797, 9.451) |
| not chronic neurological or neuromuscular disease | 0.184 (194, 1054) | Ref | 0.062 (61, 990) | Ref |
| chronic neurological or neuromuscular disease | 0.122 (6, 49) | 0.619 (0.260, 1.474) | 0.149 (7, 47) | 2.665 (1.146, 6.197) |
| not immunocompromised | 0.176 (187, 1063) | Ref | 0.060 (60, 1001) | Ref |
| immunocompromised | 0.325 (13, 40) | 2.255 (1.142, 4.453) | 0.222 (8, 36) | 4.481 (1.958, 10.255) |
| not kidney disease | 0.178 (188, 1059) | Ref | 0.058 (58, 999) | Ref |
| kidney disease | 0.273 (12, 44) | 1.737 (0.878, 3.436) | 0.263 (10, 38) | 5.794 (2.685, 12.504) |
| not current smoker | 0.182 (191, 1051) | Ref | 0.061 (60, 989) | Ref |
| current smoker | 0.173 (9, 52) | 0.942 (0.452, 1.966) | 0.167 (8, 48) | 3.097 (1.388, 6.911) |
| not obesity | 0.216 (75, 347) | Ref | 0.077 (25, 326) | Ref |
| obesity | 0.263 (65, 247) | 1.295 (0.884, 1.897) | 0.054 (12, 222) | 0.688 (0.338, 1.400) |
| unknown obesity | 0.118 (60, 509) | 0.485 (0.334, 0.703) | 0.063 (31, 489) | 0.815 (0.472, 1.408) |
| Oslo | 0.231 (80, 346) | Ref | 0.047 (15, 322) | Ref |
| Agder | 0.074 (2, 27) | 0.266 (0.062, 1.147) | 0.040 (1, 25) | 0.853 (0.108, 6.734) |
| Innlandet | 0.226 (12, 53) | 0.973 (0.488, 1.940) | 0.125 (6, 48) | 2.924 (1.075, 7.949) |
| Møre Og Romsdal | 0.313 (5, 16) | 1.511 (0.510, 4.479) | 0.071 (1, 14) | 1.574 (0.193, 12.843) |
| Nordland | 0.000 (0, 5) | 0.000 (0.000, Inf.000) | 0.000 (0, 5) | 0.000 (0.000, Inf.000) |
| Rogaland | 0.133 (10, 75) | 0.512 (0.251, 1.042) | 0.085 (6, 71) | 1.889 (0.706, 5.053) |
| Trøndelag | 0.214 (3, 14) | 0.907 (0.247, 3.330) | 0.000 (0, 13) | 0.000 (0.000, Inf.000) |
| Vestfold Og Telemark | 0.141 (9, 64) | 0.544 (0.258, 1.149) | 0.081 (5, 62) | 1.795 (0.628, 5.135) |
| Vestland | 0.282 (20, 71) | 1.304 (0.734, 2.316) | 0.031 (2, 64) | 0.660 (0.147, 2.960) |
| Viken | 0.130 (54, 415) | 0.497 (0.340, 0.727) | 0.080 (32, 398) | 1.789 (0.951, 3.366) |
| Troms Og Finnmark | 0.167 (1, 6) | 0.665 (0.077, 5.775) | 0.000 (0, 5) | 0.000 (0.000, Inf.000) |
| unknown county | 0.364 (4, 11) | 1.900 (0.542, 6.656) | 0.000 (0, 10) | 0.000 (0.000, Inf.000) |
| South-east Health Authority | 0.175 (160, 912) | Ref | 0.068 (59, 862) | Ref |
| West Health Authority | 0.207 (31, 150) | 1.224 (0.796, 1.883) | 0.058 (8, 138) | 0.838 (0.391, 1.793) |
| Mid-Norway Health Authority | 0.276 (8, 29) | 1.790 (0.779, 4.114) | 0.038 (1, 26) | 0.544 (0.072, 4.088) |
| North Health Authority | 0.083 (1, 12) | 0.427 (0.055, 3.330) | 0.000 (0, 11) | 0.000 (0.000, Inf.000) |
| COVID-19 main cause of hospitalisation | 0.200 (188, 942) | Ref | 0.057 (50, 877) | Ref |
| COVID-19 not main cause of hospitalisation | 0.071 (11, 154) | 0.309 (0.164, 0.581) | 0.118 (18, 153) | 2.205 (1.249, 3.894) |
| unknown main cause | 0.143 (1, 7) | 0.668 (0.080, 5.586) | 0.000 (0, 7) | 0.000 (0.000, Inf.000) |
| No ICU admission | NA | NA | 0.040 (35, 884) | Ref |
| ICU admission | NA | NA | 0.216 (33, 153) | 6.671 (3.996, 11.137) |

*Table 4. Adjusted hazard ratios (survival analysis) and adjusted odds ratios (logistic regression) for patient trajectories, hospitalised SARS-CoV-2 positive patients, Norway, 21 December 2020 – 25 April 2021.*

|  | HR.PreSykehus | HR.KunSykehus | HR.PreICU | HR.ICU | HR.postICU | OR, ICU | OR, death |
| --- | --- | --- | --- | --- | --- | --- | --- |
| N | 538 | 903 | 200 | 200 | 161 | 1103 | 1037 |
| events | 538 | 874 | 200 | 161 | 151 | NA | NA |
| mean time (size) /prob | 8.105 (7.825) | 4.992 (2.325) | 2.382 (1.169) | 16.826 (1.271) | 7.449 (2.139) | 0.181 | 0.062 |
| B.1.1.7 | 1.205 (0.940, 1.545) | 0.961 (0.792, 1.167) | 1.029 (0.665, 1.592) | 0.827 (0.509, 1.343) | 0.996 (0.608, 1.632) | 1.367 (0.857, 2.255) | 1.392 (0.682, 3.009) |
| age | NA (NA, NA) | 0.982 (0.978, 0.986) | 0.976 (0.966, 0.987) | 0.978 (0.965, 0.990) | 0.983 (0.972, 0.995) | 1.027 (1.017, 1.038) | 1.106 (1.079, 1.137) |
| female | NA | 1.276 (1.114, 1.462) | NA | NA | NA | NA | NA |
| cancer | NA | 0.627 (0.440, 0.895) | 0.281 (0.128, 0.616) | NA | NA | NA | 4.585 (1.907, 10.662) |
| heart disease | 1.359 (1.097, 1.684) | NA | NA | NA | NA | NA | NA |
| immunocompromised | NA | NA | NA | NA | NA | 2.328 (1.116, 4.650) | 4.146 (1.451, 11.033) |
| kidney disease | NA | 0.517 (0.343, 0.777) | NA | NA | NA | NA | NA |
| liver disease | 4.450 (1.614, 12.269) | NA | NA | NA | NA | NA | NA |
| neuro | 1.880 (1.219, 2.901) | NA | NA | NA | NA | NA | NA |
| current smoker | 2.033 (1.254, 3.294) | NA | NA | NA | NA | NA | 4.428 (1.582, 11.539) |
| obesity | NA | NA | NA | NA | NA | 1.457 (0.984, 2.159) | NA |
| unknown obesity | NA | NA | NA | NA | NA | 0.491 (0.336, 0.715) | NA |
| pregnancy | NA | NA | NA | 3.025 (1.117, 8.189) | NA | NA | NA |
| male (related to pregnancy) | NA | NA | NA | 1.140 (0.815, 1.595) | NA | NA | NA |
| ICU admission | NA | NA | NA | NA | NA | NA | 7.142 (3.942, 13.140) |
| Mid-Norway Health Authority | NA | 0.961 (0.621, 1.489) | NA | NA | 0.831 (0.365, 1.891) | NA | NA |
| West Health Authority | NA | 1.392 (1.143, 1.696) | NA | NA | 0.526 (0.319, 0.869) | NA | NA |
| North Health Authority | NA | 1.165 (0.642, 2.115) | NA | NA | 0.831 (0.365, 1.891) | NA | NA |
| Agder | 0.360 (0.196, 0.661) | NA | NA | NA | NA | NA | NA |
| Innlandet | 0.579 (0.380, 0.884) | NA | NA | NA | NA | NA | NA |
| Møre og Romsdal | 2.789 (1.107, 7.027) | NA | NA | NA | NA | NA | NA |
| Nordland | 0.997 (0.506, 1.963) | NA | NA | NA | NA | NA | NA |
| Rogaland | 0.576 (0.390, 0.853) | NA | NA | NA | NA | NA | NA |
| Trøndelag | 0.686 (0.326, 1.443) | NA | NA | NA | NA | NA | NA |
| Vestfold | 0.481 (0.306, 0.755) | NA | NA | NA | NA | NA | NA |
| Vestland | 0.548 (0.365, 0.821) | NA | NA | NA | NA | NA | NA |
| Viken | 0.590 (0.441, 0.790) | NA | NA | NA | NA | NA | NA |
| unknown county | 1.241 (0.385, 3.996) | NA | NA | NA | NA | NA | NA |
